# Duration of SARS-CoV-2 viral shedding in faeces as a parameter for wastewater-based epidemiology: Re-analysis of patient data using a shedding dynamics model

**DOI:** 10.1101/2020.11.22.20236323

**Authors:** Fuminari Miura, Masaaki Kitajima, Ryosuke Omori

## Abstract

**Background:** Wastewater-based epidemiology (WBE) is one of the most promising approaches to effectively monitor the spread of the novel coronavirus disease 2019 (COVID-19). The virus concentration in faeces and its temporal variations are essential information for WBE. While some clinical studies have reported severe acute respiratory syndrome coronavirus 2 (SARS-CoV-2) concentrations in faeces, the value varies amongst patients and changes over time.

**Aim:** The present study aimed to examine how the temporal variations in the concentration of virus in faeces affect the monitoring of disease incidence. We re-analysed the experimental findings of clinical studies to estimate the duration of virus shedding and the faecal virus concentration.

**Method:** Available experimental data as of 23 October, 2020 were collected and patient data reported in Germany were included for further analysis. The viral shedding kinetics was modelled, and the dynamic model was fitted to the collected experimental data by a Bayesian framework. Using samples of posterior distributions, the duration of viral shedding and the concentration of virus copies in faeces over time were computed.

**Results:** We estimated the median concentration of SARS-CoV-2 in faeces as 2.6 (95% Credible Interval (CrI): 0.22–4.8) log copies per gram (g) of faeces over the shedding period, and our model implied that the duration of viral shedding was 23.2 days (95% CrI: 19.5–31.5), given the current standard quantification limit (Ct = 40). With simulated incidences, our results also indicated that a one-week delay between symptom onset and wastewater sampling increased the estimation of incidence by 13.5%.

**Conclusions:** Our results demonstrated that the temporal variation in virus concentration in faeces affects microbial monitoring systems such as WBE. The present study also implied the need for adjusting the estimates of virus concentration in faeces by incorporating the kinetics of unobserved concentrations. The method used in this study is easily implemented in further simulations; therefore, the results of this study might contribute to enhancing disease surveillance and risk assessments that require quantities of virus to be excreted into the environment.

## 1. Introduction

As of 23 October 2020, the novel coronavirus disease 2019 (COVID-19) has spread all over the world. Severe acute respiratory syndrome coronavirus 2 (SARS-CoV-2) is the causative agent of this disease. The infection causes general symptoms such as fever, cough, shortness of breath and diarrhoea [1], and the disease progression differs according to age [2,3] and clinical history [4]. Previous epidemiological studies on COVID-19 have reported that a substantial proportion of infected individuals were even asymptomatic [5,6]. These disease characteristics have caused complications in controlling the transmission of SARS-CoV-2.

To identify hidden chains of infection, more effective ways of monitoring the spread of the disease are required. In this context, wastewater-based epidemiology (WBE) has been attracting attention as a promising potential approach [7,8]. Clinical observations have demonstrated that there is prolonged virus shedding in faeces, ranging from 1 to 33 days [9–11], and therefore wastewater surveillance can enable the monitoring of excreted viruses through which we can capture the presence and possibly the numbers of infected individuals, regardless of their symptoms. As a monitoring system, researchers have successfully reported their detection or quantification results in various countries, such as Australia [12], Japan[13], etc. [14–16]. The Netherlands and several municipalities in the United States have already started utilising WBE in practice as a part of their surveillance systems [17,18].

One of the most important quantities for WBE is the SARS-CoV-2 concentration in faeces. If we wish to simulate the potential number of cases in a sewershed, the virus concentration in faeces and its time course would affect the simulation. For example, earlier WBE studies have attempted to simulate the incidence using sewage data [12,19]; however, no previous studies have accounted for temporal variations in the virus concentration in faeces, and thus undetectable concentrations that might occur after several weeks from symptom onset have not been considered. In addition, while several clinical studies have reported that there is prolonged SARS-CoV-2 shedding in faeces with binary results (i.e. positive/negative) [11] or threshold cycle (Ct) values over time [10,20], quantitative estimates of possible virus excretions have yet to be fully explored.

The present study aims to examine how temporal variations in virus concentrations in faeces affect disease incidence monitoring systems such as WBE by estimating the duration of virus shedding and the virus concentration in faeces. We screened available time course data of virus concentration in faeces in clinical studies and subsequently applied a mathematical model that describes the kinetic viral shedding process. The present study also simulated the potential bias for the estimation of incidence with WBE by using an estimated virus shedding time course.

## 2. Methods

### 2.1 Data collection

A literature review was conducted to collect available experimental data as of 23 October, 2020 using Google Scholar, PubMed and MedRxiv. Seven articles containing the following information were assessed with full text reviews: (a) exact virus copies or Ct values, (b) days from symptom onset or days from hospitalisation and (c) the amount of stool that was quantified by quantitative polymerase chain reaction (qPCR) [10,20–25]. As a result of this review, only patient data reported in Wolfel et al. (2020) were used for further analysis [21]; the other studies were excluded because they reported only Ct values and the experimental details required conversion to virus concentration in faeces [copies/grams (g)-faeces].

The analysed data consist of hospitalised patients involved in a large cluster that occurred in Munich, Germany between 23 and 27 January, 2020 [21]. Since it is difficult in retrospective studies to collect stool samples before the onset of symptoms, the data were expressed in the unit of time ‘days after symptom onset’ [21]. The sample size reported in this study was insufficient to stratify the study population into sub-groups and thus exhaustive data were analysed to estimate the temporal variations in virus shedding.

### 2.2 Model

The time course of virus shedding is modelled as two processes. First, the virus is accumulated in a human host with concentration *c*_*1*_(*t*) and then, at a certain point, the virus is shed in concentration *c*_*2*_(*t*). According to Teunis et al. [26], the dynamics can be formulated simply as follows:

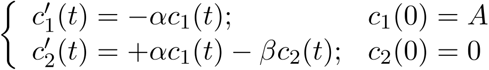

where A is the initial concentration of viruses and *α* and *β* are the transport rates of viruses. By solving this system, the time course of virus concentration in faeces can be written as

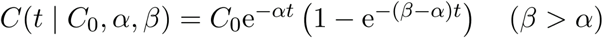

where 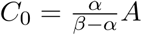 (see a more detailed biological description in the original study [26]). In the present study,*t* = 0 was defined as the day of symptom onset, assuming that the accumulation of viruses was complete when the individuals started shedding viruses in their faeces.

One of the properties of this model is that one can analytically calculate important quantities such as peak concentration. Finding the extrema of *C*(*t*), the time to peak concentration *t*_*peak*_ is written as 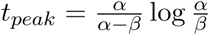, and the peak concentration *C*_*peak*_ is consequently obtained as 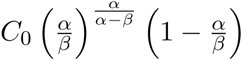.

To define the end of virus shedding, Ct = 40 was used as the threshold (corresponding to approximately 10^2^ copies per g-faeces), following the recommended quantification threshold value in current qPCR protocols [21]. The duration of virus shedding *t*_*total*_ was then computed by seeking the intersection point of *C*(*t*) and the threshold value.

### 2.3 Model fit

To jointly estimate the parameters (*C*_*0*_, *α, β*), a hierarchical Bayesian framework was used to account for individual variations and provide for the uncertainty of the estimates. Here, we denote the expected value as *𝒰* = In(*C*(*t*)) with natural log scale. By assuming that the log-transformed concentration is normally distributed, the likelihood is simply

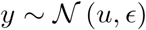

where *𝒴* is the observed data (i.e. the log-scaled virus concentration data) and *ϵ* is the standard deviation. We describe the variation in the observed data in a hierarchical model framework

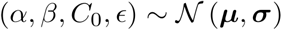

with mean vector 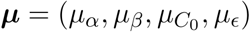 and standard deviation vector 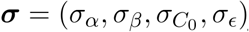 .

For the hyperparameters, we specified positive non-informative prior probability distributions for parameters *μ* and *σ* Using the Hamiltonian Monte Carlo (HMC) algorithm, we obtained estimates and 95% credible intervals (CrI) by sampling the posterior distributions. All of the computations above were implemented in R-4.0.0 with a package {rstan}-2.21.2. [27,28].

### 2.4 Simulation of relative incidence

Suppose that we wish to calculate the potential number of infected individuals per day using WBE surveillance. If we have observational data on the average number of SARS-CoV-2 ribonucleic acid (RNA) copies in sewage per day 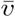 and complete information on the symptom onset of all of the infected individuals, we can simulate the crude daily incidence *Ī* with the average daily water volume 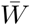 and the average amount of faeces per person per day 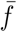 :

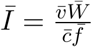

where 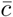 is the average virus concentration in faeces.

The crucial point here is that the virus concentration is the variable that is dependent on the time between sewage sampling and symptom onset. We thus denote the concentration in faeces at day *τ* from symptom onset as 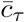 and examine the sensitivity of the expected incidence to the virus concentration in faeces by defining the relative incidence as

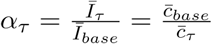

Where 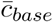 is the baseline concentration defined by the median of the (quantified) concentration reported in Wolfel et al. [21]. For simplicity, the analysis assumes that all of the individuals had the same symptom onset (days before sampling) and we simulated *τ* = 1, 7, 14, 21, 28 by changing 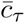 and fixing the other conditions.

## 3. Results

### 3.1 Time course of viral shedding

The estimated curve for the time course of virus shedding is shown in **Fig-1**. The results of model fitting indicate that the virus concentration in faeces rapidly increases after symptom onset, and that prolonged shedding occurs until the concentration becomes lower than the quantification limits. The time to peak concentration *t*_*peak*_ was estimated to be 0.38 (95% CrI: 0.22–2.6) days. If we define the end of virus shedding as the time point at which the virus concentration becomes lower than the quantification limit of qPCR (in this analysis, two log(copies)), the duration of virus shedding was estimated to be 23.2 (95% CrI: 19.5–31.5) days.

**Fig-1.**
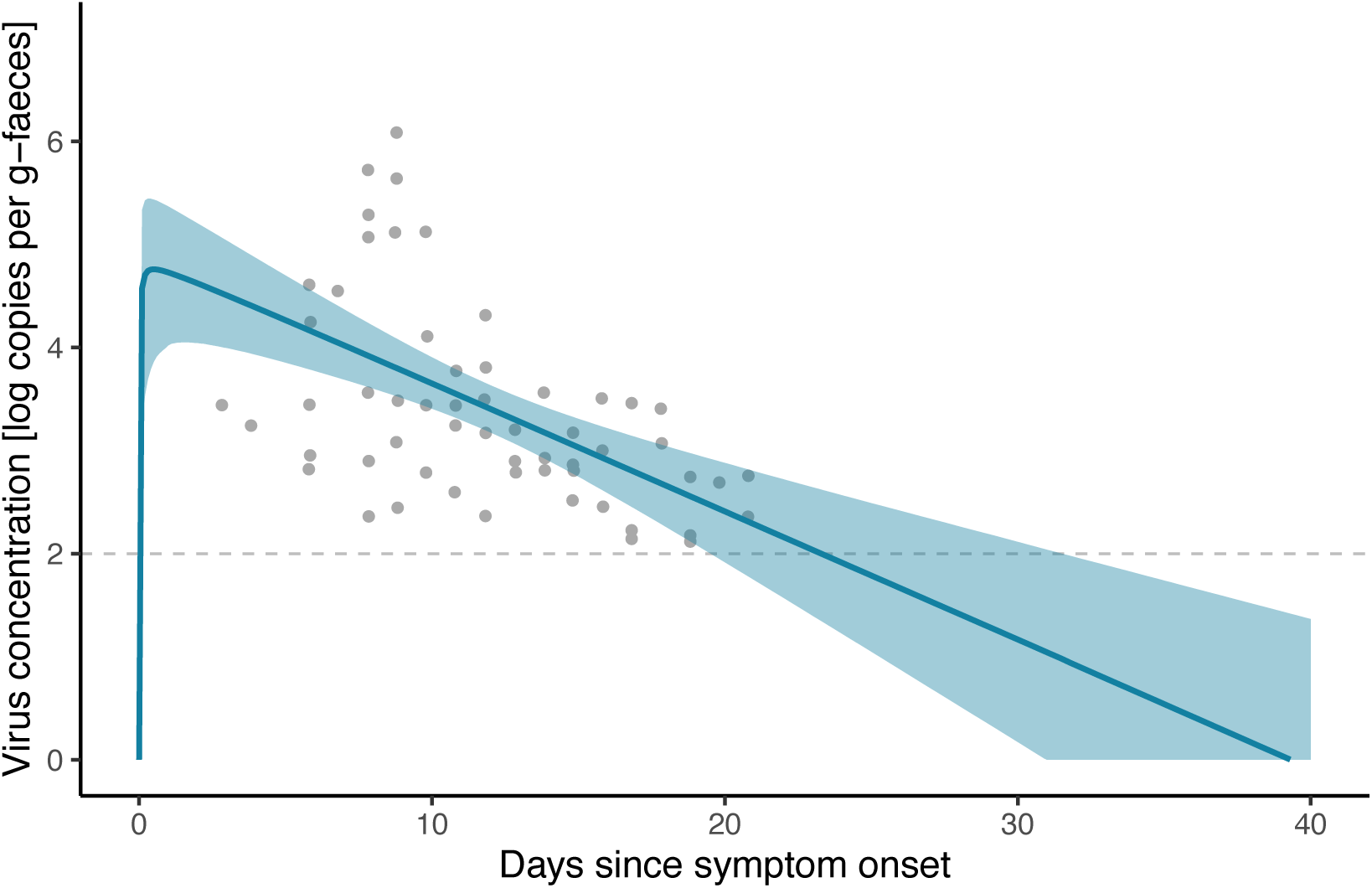
Fitted model to the observed virus concentration in faeces. Grey plots represent observed data, the blue curve shows the posterior median and the shaded zones are the 95% credible intervals. The dashed line indicates the quantification limit threshold of qPCR (2 log copies/g-faeces).

### 3.2 Expected virus concentration

With HMC posterior samples, we estimated the possible range of virus concentrations in faeces during the shedding period and at peak concentration. Those estimates with uncertainty are shown in **Fig-2(A)**. If we use the quantitative data from Wolfel et al. to calculate the expected virus concentration [21], the median was 3.2 (95% percentile: 2.2–5.7) [log copies per g-faeces]; however, our estimates suggest that the median of concentration over the whole shedding period was 2.6 (95% CrI: 0.22–4.8) [log copies per g-faeces]. For peak concentration *C*_*peak*_, the estimate was 4.8 (95% CrI: 4.1–5.5) [log copies per g-faeces].

**Fig-2.**
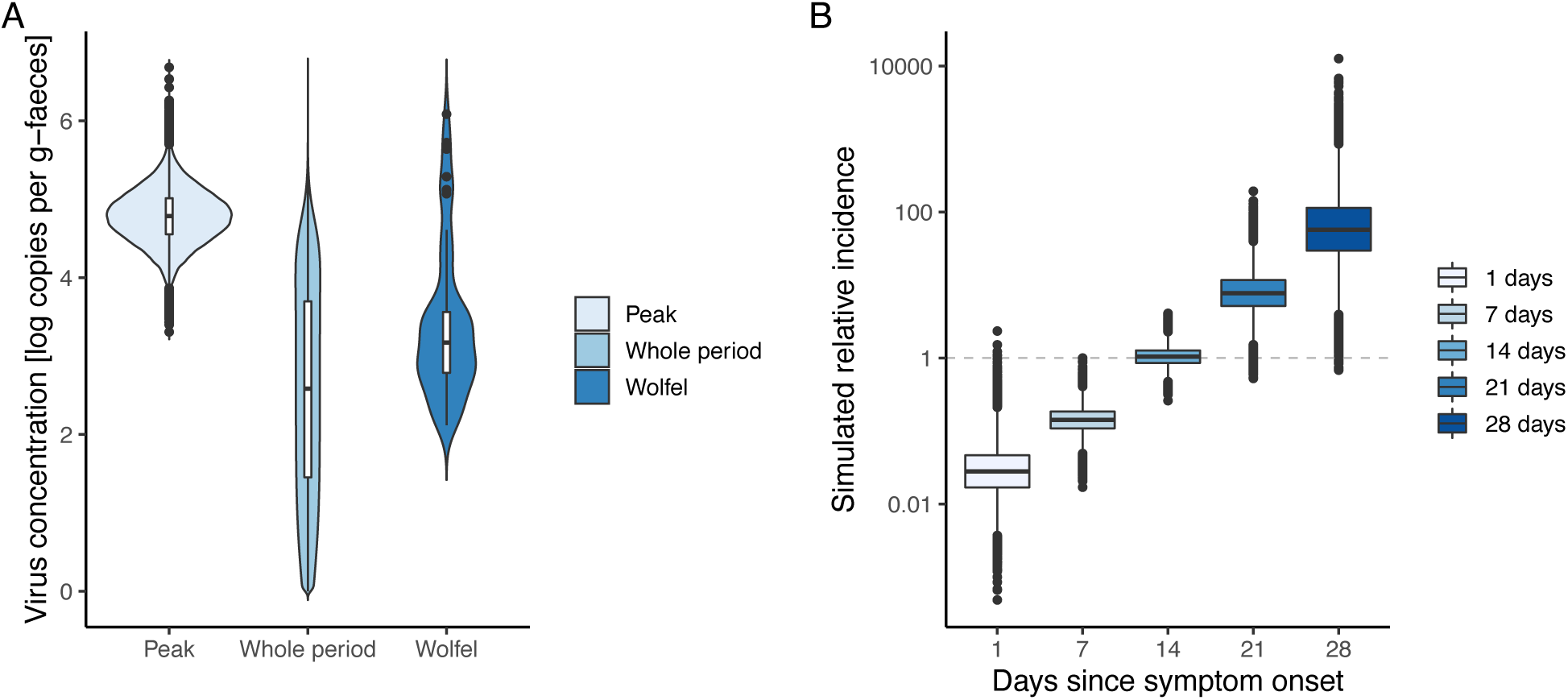
Estimated virus concentration in faeces **(A)** and simulated relative incidence caused by changing the delay between wastewater sampling and symptom onset **(B). (A)** From the left of the panel, violin plots of estimated concentrations in faeces on the day of peak shedding, of the entire shedding period and of raw quantified experimental data reported by Wolfel et al. are visualised [21]. **(B)** Relative incidences for different sewage samples are plotted, in which we assumed that all of the individuals had the same symptom onset (days before sampling, as shown on the x-axis). By definition, the relative incidence equals 1 when the quantitative data in Wolfel et al. are used for the virus concentration in faeces [21].

### 3.3 Simulated relative incidence

To assess the potential impact of the timing of sewage sampling for WBE, we simulated the relative incidence by changing the delay between sewage sampling and symptom onset. **Fig-2(B)** illustrates the results of simulated relative incidences in which the value equals 1 when Wolfel et al.’s quantified data are used for the virus concentration in faeces [21]. The results indicate that the (log-converted) expected incidence increased linearly with longer intervals, and a one-week delay increased the estimated incidence by 13.5% (i.e., 10^0.87^ times higher).

## 4. Discussion

In this study, we estimated the time course of SARS-CoV-2 shedding in faeces by applying a simple dynamic model. Our findings show that there is a large temporal variation in virus concentration in faeces (**Fig-1**), resulting in potential bias when simulating the number of infected cases with the data on virus concentration in sewage (**Fig-2**). The model describes the kinetics of the accumulation and excretion processes (**Eq-1**) and thereby accounts for unobserved concentrations that were under the detection limits of real-time PCR.

The duration of virus shedding in faeces was estimated to be 23.2 (95%CrI: 19.5–31.5) days from symptom onset. This result is consistent with previous clinical reports [9,11]. While earlier studies have suggested possible durations with only binary (positive/negative) results [11], our method provides a credible range of shedding duration with uncertainty. Furthermore, we can easily simulate it even when the threshold values are changed. Several clinical studies have reported Ct values over time, but the amount of stool in RNA extractions was not recorded [20,22,23,25]. If such information is available, that evidence would be synthesised with our method and induce more valuable implications for both the natural history of COVID-19 infection and environmental surveillance.

In the context of WBE, our findings on the peak and median concentrations indicated that there might be bias if we use only quantified experimental results of faecal samples by truncating unobserved concentrations. There are large variations in concentration depending on the timing of taking faecal samples (**Fig-1** and **Fig-2(A)**); a one-week delay increases the estimation of incidence by 13.5% (**Fig-2(B)**). Previous studies have attempted to simulate the potential incidence with the quantified virus concentration in sewage [12,19]; however, since there is a strong dependency between the observed concentration in sewage and the sampling delay from symptom onset, we must carefully specify the interpretation of quantities averaged over time. Ideally, epidemiological data such as case series based on symptom onset should be incorporated to simulate incidences more precisely.

There are several limitations in this study. First, the analysed data are a single cohort that consists of diagnosed cases in Germany. The disease progression might differ by age, race or medical history and, consequently, those factors might affect the estimates. Since the sample size was not sufficient to stratify the data into sub-groups, additional data are needed for further analysis. Second, our modelling assumed that the virus shedding in faeces starts from symptom onset. If the peak had occurred before symptom onset, it would not be captured with this analysis. While earlier clinical studies have indicated that the peak in faeces might be around symptom onset [10,20] and in throat swabs [29], there was no publicly available data that contain viral loads from the day of infection to symptom onset. To obtain a conclusive estimate of the peak timing, we need observational data such as (prospective) periodic stool sampling or human challenge studies.

Despite the abovementioned limitations, our analysis would be beneficial for surveillance systems in different sectors. Our findings indicate that the temporal variation in virus concentrations affects microbial monitoring systems such as WBE and repeated testing in hospitals. Especially for the estimation of incidence, the virus concentrations in faeces must be adjusted by incorporating the kinetics of unobserved concentrations (i.e. concentrations lower than the quantification limits). The method used in this study is easily implemented in other simulations and therefore the results of this study might contribute to enhancing disease surveillance and risk assessments that require data on the quantities of viruses that have been excreted into the environment.

### CRediT author contribution statement

Fuminari Miura: Conceptualization, Methodology, Data curation, Formal analysis, Writing - Original draft, Masaaki Kitajima: Writing - Review & Editing, Funding acquisition, Ryosuke Omori: Methodology, Writing - Original draft, Funding acquisition

## Data Availability

All relevant data are within the manuscript.

## Acknowledgment

The authors would like to thank Enago for the English language editing.

